# The HIV and STI syndemic following mass scale-up of combination HIV interventions in Uganda: a population-based cross-sectional study

**DOI:** 10.1101/2022.03.31.22273254

**Authors:** M. Kate Grabowski, Josephine Mpagazi, Stephen Kiboneka, Robert Ssekubugu, John Baptiste Kereba, Annet Nakayijja, Julius Tukundane, Jade Jackson, Austin D. Peer, Caitlin Kennedy, Godfrey Kigozi, Ronald M. Galiwango, Yuka Manabe, Larry W. Chang, Sarah Kalibala, Ronald H Gray, Maria J Wawer, Steven J Reynolds, Aaron AR Tobian, David Serwadda, Charlotte A. Gaydos, Joseph Kagaayi, Thomas C Quinn, the Rakai Health Sciences Program

## Abstract

**Background:** Combination HIV interventions (CHIs) have led to significant declines in HIV incidence in sub-Saharan Africa; however, population-level data on non-HIV sexually transmitted infection (STI) burden in the context of CHIs are rare. We aimed to assess STI burden in Uganda following mass scale-up of CHIs, including universal HIV treatment.

**Methods:** The Sexually Transmitted Infection Prevalence Study (STIPS) was a cross-sectional study nested within the Rakai Community Cohort Study (RCCS), a population-based cohort among inland agrarian and Lake Victoria fishing populations in southern Uganda. STIPS enrolled consenting participants, 18-49 years, between May and October 2019 and measured prevalence of *Chlamydia trachomatis* (chlamydia), *Neisseria gonorrhoeae* (gonorrhea), *Trichomonas vaginalis* (trichomonas), syphilis, and herpes simplex virus type 2 (HSV-2).

**Findings:** STIPS enrolled 1,825 participants, including 965 women (53%), of whom 9% (n=107) were pregnant. Overall, there was 9.8% prevalence of chlamydia (95%CI:8.5-11%), 6.7% gonorrhea (95%CI:5.7-8.0%), and 11% trichomonas (95%CI:9.5-12%). In the fishing population, syphilis reactivity was 24% (95%CI:22-27%), with 9.4% (95%CI:7.7-11%) having high titer (RPR ≥ 1:8) infection, including 17% (95%CI:12-24%) of HIV-positive men. Prevalence of ≥ 1 curable STI (chlamydia, gonorrhea, trichomonas, or high titer syphilis) was 44% higher among HIV-positive persons (adjusted prevalence risk ratio [adjPRR]=1.44,95%CI:1.22-1.71), with no differences by HIV treatment status. HIV-positive pregnant women were more likely than HIV-negative pregnant women to have a curable STI (adjPRR=1.87, 95%CI: 1.08-3.23).

**Interpretation:** STI burden remains extremely high in Uganda, particularly among HIV-positive persons. There is an urgent need to integrate STI diagnostic testing and treatment with HIV services in African settings.

**Funding:** National Institutes of Health

## INTRODUCTION

Sexually transmitted infections (STIs) are a major cause of fetal and neonatal deaths, cervical cancer, and infertility, and are associated with reduced quality of life.^1^ Globally, the absolute number of STI cases and associated disability adjusted life years (DALYS) have increased between 1990 and 2019, despite reductions in HIV incidence with mass scale-up of combination HIV prevention and treatment interventions (CHIs).^2,3^ In 2019, the US Centers for Disease Control (CDC), reported more than 2.5 million cases of Chlamydia trachomatis (chlamydia), Neisseria gonorrhea (gonorrhea), and syphilis, continuing an alarming, rapid rise in STIs since 2015.^4^ Emerging data on STI trends in high-income countries, including the US, have prompted re-evaluation of the relationship between HIV and STIs in the context of CHIs.^5^

While there are relatively abundant data on STIs in high-income countries, there are limited data on STI trends and their relationship to CHIs elsewhere. Incidence rates of chlamydia, gonorrhea, and Trichomonas vaginalis (trichomonas) are highest in sub-Saharan Africa, with the majority of cases occurring among heterosexual men and women.^6^ The region also bears the world’s greatest HIV burden with >25 million people living with HIV.^7^ Since the mid-2000s, there has been substantial global investment in African HIV epidemic control.^8^ HIV interventions, including antiretroviral therapy (ART) and voluntary medical male circumcision (VMMC), have been successfully rolled out and scaled in many countries.^9^ While data from studies across Africa show declining HIV incidence with increasing coverage of CHIs,^10^ population-level data on STI burden in the era of widespread biomedical HIV prevention is rare. Understanding the extent to which modern HIV and STI epidemics in Africa intersect could identify potential points of synergy for integrated disease control within African HIV treatment and prevention programs.^11^

Here, we report results from the Sexually Transmitted Infection Prevalence Study (STIPS), a cross-sectional, population-level study of chlamydia, gonorrhea, trichomonas, syphilis, and herpes simplex virus 2 prevalence. STIPS was nested in the Rakai Community Cohort Study (RCCS), a population-based cohort ongoing since 1994 in south-central Uganda. Scale-up of CHIs have led to significant declines in HIV incidence in RCCS study communities;^12,13^ however, the burden of STIs since CHI rollout has not been characterized previously.

## METHODS

### The Sexually Transmitted Infection Prevalence Study (STIPS)

STIPS was a cross-sectional study nested in the nineteenth survey round (R19) of the Rakai Community Cohort Study (RCCS), an open population-based census and HIV cohort study in Rakai and surrounding districts in southern Uganda conducted by the Rakai Health Sciences Program (RHSP)^14^. As part of routine activities, the RCCS conducts an annual census with no age truncation. The RCCS survey, conducted after the census, includes all consenting residents. Interviewers use structured questionnaires programmed on mobile PCs to collect sociodemographic, behavioral, and health information.

Enrollment into STIPS was restricted to RCCS participants aged 18-49 years residing in two study communities. Selected communities included a semi-urban trading center and surrounding rural villages and a fishing community along the Lake Victoria shoreline, herein referred to as inland and fishing communities, respectively. These communities were selected due to their geographic diversity, population size, and enrollment timelines within the R19 survey. RCCS participants were separately consented for STIPS, and those who agreed to participate provided written informed consent for collection of three genital swabs (for chlamydia/gonorrhea testing, for trichomonas testing, and for storage for future studies respectively). Clinician collected penile meatal swabs and self-collected vaginal swabs were obtained from men and women, respectively.

Prior RHSP research has shown that self-administered vaginal swabs are comparable to clinician-collected swabs for women with high acceptability and concordance of STI results.^15^ STIPS participants also were administered an STI module during the RCCS survey, which included survey questions on STI symptoms (current and last 6 months) and STI treatment seeking behaviors. Enrollment of STIPS participants took place between May 27, 2019 and October 25, 2019.

STIPS was approved by Uganda Virus Research Institute Research Ethics Committee, the Ugandan National Council for Science and Technology, and the Johns Hopkins School of Medicine Institutional Review Board. All study participants provided written informed consent for STIPS in addition to the RCCS.

### Procedures

HIV, HSV-2, and syphilis antibody tests were performed using sera from peripheral blood samples collected from STIPS participants. HIV and syphilis testing was done at time of survey and HSV-2 testing after enrollment of all STIPS participants. HIV testing was done using a field-validated parallel three test rapid HIV testing algorithm with enzyme-linked immunosorbent assays (ELISA) and PCR testing for first-time diagnoses ^16^. HIV viral load testing was done for all HIV seropositive participants using the Abbott RealTime assay (Abbot Molecular, Inc., Des Plaines, IL), and an HIV viral load ≥ 1000 copies per ml was classified as viremic. HSV-2 testing was performed using the Kalon ELISA (Kalon Biological Ltd, Guilford UK) with a previously validated index cutoff value of 1.5.^17^

Syphilis antibody status was determined using the SD Bioline Syphilis 3.0 (SD Biostandard Diagnostics Private Limited, Gurgaon, Haryana, India), a solid phase immunochromatographic point-of-care assay for the qualitative detection of treponemal antibodies of IgG, IgM and IgA isotype. The rapid plasma reagin test (RPR, Cypress Diagnostics, Hulshout, Belgium) was performed for all participants with positive antibody results on SD Bioline to determine syphilis titers. RPR testing was done within 24 hours at the RHSP central laboratory, and active syphilis infection was defined as an RPR titer ≥ 1:8.

Penile meatal and vaginal swabs were tested for chlamydia, gonorrhea, and trichomonas. Chlamydia and gonorrhea testing was performed using the Abbott m2000 RealTime CT/NG assay for the direct, qualitative detection of plasmid DNA of chlamydia and genomic DNA of gonorrhea. Testing was performed at the RHSP central laboratory within 7 days of sample collection according to the manufacturer’s protocol. Trichomonas testing was performed from genital swabs at time of survey using the OSOM Trichomonas Rapid Test (Sekisui [formerly Genzyme Diagnostics], Burlington, MA) for qualitative detection of T. vaginalis antigens.

If a woman self-reported that she was pregnant and her pregnancy was visually confirmed by the interviewer, she was confirmed to be currently pregnant. If the pregnancy could not be visually confirmed or she reported that she was not pregnant, she was asked to provide the date of the first day of her last menstrual period (LMP). Any woman who reported an LMP >30 days and who was not using a hormonal birth control implant was administered a lateral flow urine pregnancy test (Cypress Diagnostics, Hulshout, Belgium).

### Return of STI test results and provision of treatment

HIV, syphilis and trichomonas rapid tests results were returned to participants immediately using on-site post-test counselors. If results were positive, they were offered free treatment per the United States Centers for Disease Control and Prevention (CDC) treatment guidelines and Uganda Clinical Guidelines for treatment of STIs.^18,19^ Individuals who reported symptomatic vaginal or penile discharge or genital ulcers at time of survey but who were not diagnosed with an STI were also offered treatment in line with World Health Organization (WHO) syndromic management guidelines.^20^ Participants who tested positive for chlamydia and gonorrhea were re-contacted after the survey and were provided with post-test counseling and free treatment if they had not already been treated at time of survey

### Statistical analysis

We have previously shown that HIV prevalence significantly varies between Lake Victoria fishing and inland communities in the RCCS ^14^, and so we first compared the demographic, sexual behavior, and health service characteristics between STIPS participants residing in inland and fishing communities. All characteristics were described categorically using frequency counts and percentages, and statistically significant differences were determined using chi-square tests. We next estimated the prevalence of each STI separately by gender, community type (i.e., fishing vs. inland), and HIV status using modified Poisson regression with robust variance estimators ^21^. Prevalence was estimated similarly by age group (15-24, 25-29. 30-34. 35-39, 40-44, 45-49 years), gender, and community type. Overlap in HIV and STI epidemics was evaluated using frequency counts with the *UpSetR* package in the R statistical software (version 4.0.3).

We also analyzed risk factors for curable STIs, including chlamydia, gonorrhea, trichomonas and active syphilis. Risk factors evaluated included gender, age group, marital status, educational level, number of sex partners in the last year, HIV serostatus, male circumcision, and pregnancy. Associations between HIV viremia and pre-exposure prophylaxis for HIV prevention (PrEP) use and STIs were also evaluated but restricted to HIV-positive and negative participants, respectively. All associations were measured using unadjusted and adjusted modified Poisson regression and reported as unadjusted and adjusted prevalence risk ratios (PRR) with 95% confidence intervals (95%CI).

HIV incident cases were defined as participants with a first positive HIV test result in the STIPS/RCCS R19 survey and a prior HIV-negative test result at the previous RCCS survey (R18) allowing for up to one missed visit (R17 if missed R18) as previously described ^12^. Associations between STIs and HIV incidence were measured using Poisson regression models and reported as incidence rate ratios (IRR) with 95% CI.

Lastly, we measured the validity of self-reported current STI symptoms for diagnosis of chlamydia, gonorrhea, and trichomonas (i.e., syndromic STI management). Clinically indicative symptoms of each STI were defined as specified in the 2016 Uganda clinical guidelines ^19^. Sensitivity and specificity of each self-reported symptom, clinically indicative symptoms, and all symptoms combined were estimated using the *epiR* package in R.

## RESULTS

### Study participation and characteristics of the study population

There were 2,596 census eligible participants, of whom 1,890 (73%) were present in the community at time of the RCCS survey. Of those individuals present, 1,824 (97%) consented to participate in STIPS, including 919 participants in the inland community and 906 in the fishing community (Table 1). The median age of study participants was 32 years (IQR: 25-39) and 965 (53%) were women of whom 107 were pregnant. HIV prevalence in inland and fishing communities was 14 and 40%, respectively, with ∼90% HIV viral load suppression in both communities. Half of men were circumcised (n=430/860), but few participants reported ever using PrEP (n=43, 2.4%). Fishing community participants were significantly more likely to be male, older, previously married, less likely to be educated, and to have greater numbers of sexual partners compared to inland community participants (Table 1).

**Table 1.** Characteristics of STIPS participants by community type

|  | **Inland** | **Fishing** | **p-value\*** |
| --- | --- | --- | --- |
|  | N (%) | N (%) |  |
| **Overall** | 919 | 906 |  |
| **Sex** |  |  | p=0.0009 |
| Female | 522 (56.8) | 443 (48.9) |  |
| Male | 397 (43.2) | 463 (51.1) |  |
| **Age** |  |  |  |
| 15-19 | 262 (28.6) | 177 (19.5) | p=0.0001 |
| 25-29 | 156 (17.0) | 192 (21.2) |  |
| 30-34 | 136 (14.8) | 169 (18.7) |  |
| 35-39 | 162 (17.6) | 170 (18.8) |  |
| 40-44 | 123 (13.4) | 131 (14.5) |  |
| 45-49 | 80 (8.7) | 67 (7.4) |  |
| **Marital Status** |  |  |  |
| Currently married | 605 (65.8) | 595 (65.7) | p<0.0001 |
| Never married | 169 (18.4) | 93 (10.3) |  |
| Previously married | 145 (15.8) | 218 (24.1) |  |
| **Education** |  |  |  |
| None | 36 (3.9) | 97 (10.7) | p<0.0001 |
| Primary | 537 (58.4) | 607 (67.0) |  |
| Secondary | 251 (27.3) | 154 (17.0) |  |
| Tertiary/Trade School | 95 (10.3) | 48 (5.3) |  |
| **Occupation** |  |  | p<0.0001 |
| Agriculture | 482 (52.3) | 35 (3.9) |  |
| Transport | 31 (3.4) | 23 (2.5) |  |
| Fishing | 4 (0.4) | 285 (31.5) |  |
| Government/clerical/teaching | 60 (6.5) | 11 (1.2) |  |
| Hair dresser | 34 (3.7) | 19 (2.1) |  |
| Housework | 22 (2.4) | 89 (9.8) |  |
| Student | 44 (4.8) | 8 (0.8) |  |
| Trader/shop keeper | 131 (14.3) | 204 (22.5) |  |
| Bartender/waitress | 16 (1.7) | 76 (8.4) |  |
| **Sex partners in the last year** |  |  | p<0.0001 |
| None | 117 (12.7) | 62 (6.8) |  |
| One | 571 (62.1) | 485 (53.5) |  |
| Two | 149 (16.2) | 190 (21.0) |  |
| Three | 52 (5.7) | 93 (10.3) |  |
| Four or more | 39 (3.3) | 62 (6.8) |  |
| **HIV prevalence** |  |  | p<0.0001 |
| HIV negative | 790 (86.0) | 545 (60.2) |  |
| HIV positive | 129 (14.0) | 361 (39.8) |  |
| **Viral load suppression status** |  |  | p=0.83 |
| ≤1000 copies/ml | 118 (91.2) | 326 (90.3) |  |
| >1000 copies/ml | 11 (8.5) | 35 (9.7) |  |
| **Male circumcision status\*\*** |  |  | p=0.41 |
| Uncircumcised | 205 (51.6) | 225 (48.6) |  |
| Circumcised | 192 (48.4) | 238 (51.4) |  |
| **Ever used PrEP\*** |  |  |  |
| No | 785 (99.4) | 505 (92.7) | p<0.0001 |
| Yes | 4 (0.5) | 39 (7.2) |  |
| **Currently pregnant** |  |  | p=1 |
| No | 464 (88.9) | 394 (88.9) |  |
| Yes | 58 (11.1) | 49 (11.1) |  |

\*There was missing data on PrEP use for 2 participants \*\*Male circumcision status was not clinically confirmed for 7 male participants (5 self-reported being uncircumcised and 2 circumcised).

### Prevalence of STIs by community, age, sex, and HIV serostatus

There were a total of 593 chlamydia (n=177), gonorrhea (n=122), trichomonas (n=196), and active syphilis infections detected in 478 (26%) study participants. HIV-positive participants were 56% more likely to diagnosed with ≥ 1 of these curable STIs compared to HIV-negative participants (36% vs. 23%; adjPRR=1.44, 95%CI:1.22-1.71). Figure 1 shows overlap of HIV and curable STI epidemics. The most frequently co-occurring infection with HIV was trichomonas followed by active syphilis and then gonorrhea.

**Figure 1.**
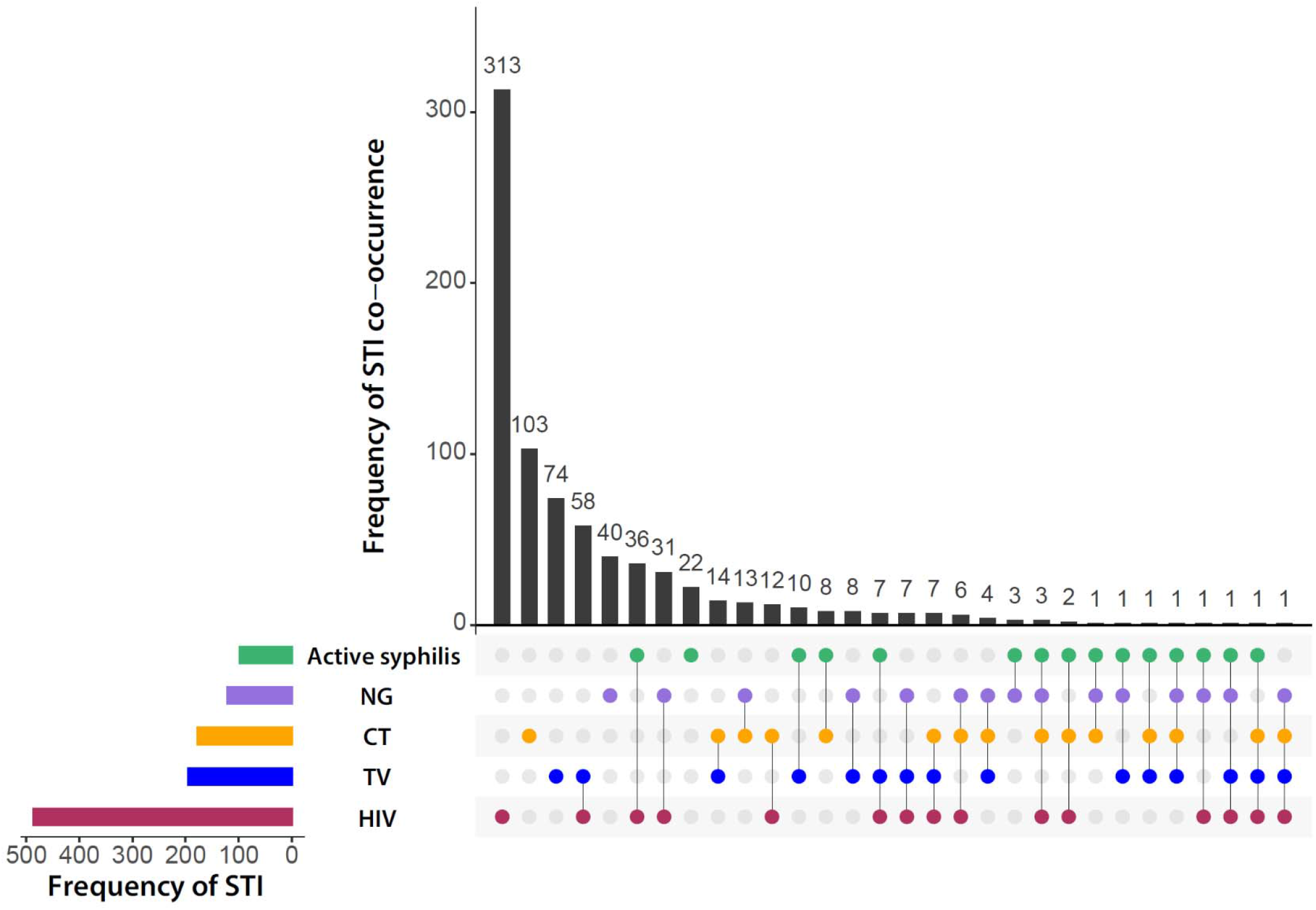
Frequency of co-occurrence of HIV, active syphilis, gonorrhea (NG), chlamydia (CT), and trichomoniasis (TV) among 490 HIV-positive and 1335 HIV-negative STIPS participants.

Table 2 shows prevalence of STIs among men and women in each community stratified by gender and HIV serostatus. Overall, chlamydia prevalence was 9.8% (n=177/1815; 95%CI: 8.5-11.2). While chlamydia levels did not differ between participants by community or gender, prevalence tended to be lower among HIV-positive compared to HIV-negative participants (6.6 vs 10.9%, adjPRR=0.75. 95%CI: 0.52-1.09) (Supplemental Table 1). Among men and women, chlamydia prevalence was highest among those <30 years, and significantly declined with increasing age. Age-specific trends of chlamydia prevalence were similar between inland and fishing communities (Figure 2A and B).

**Table 2:**
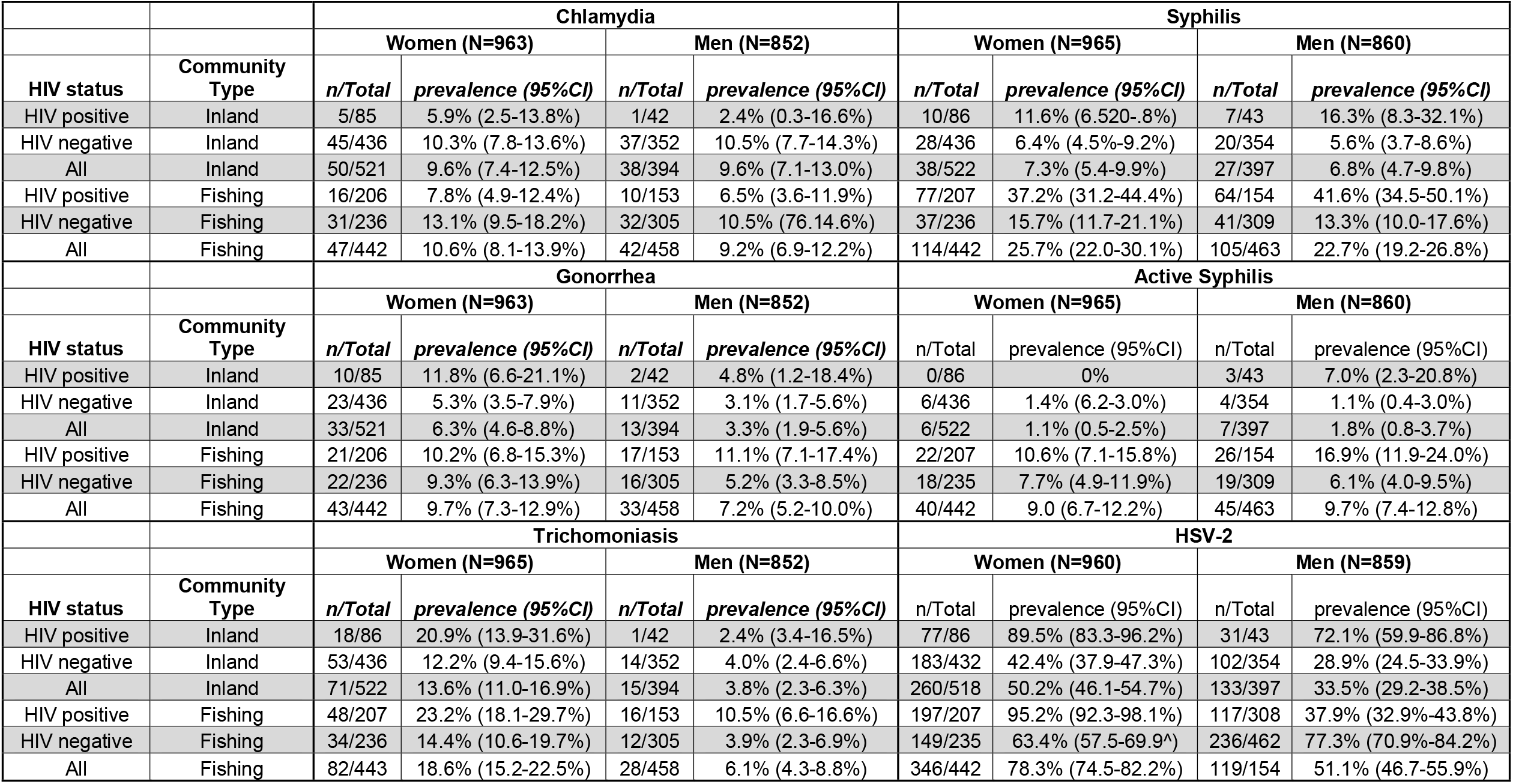
Prevalence of sexually transmitted infections among HIV-positive and HIV-negative men and women by community type (agrarian, trading, fishing).

**Figure 2.**
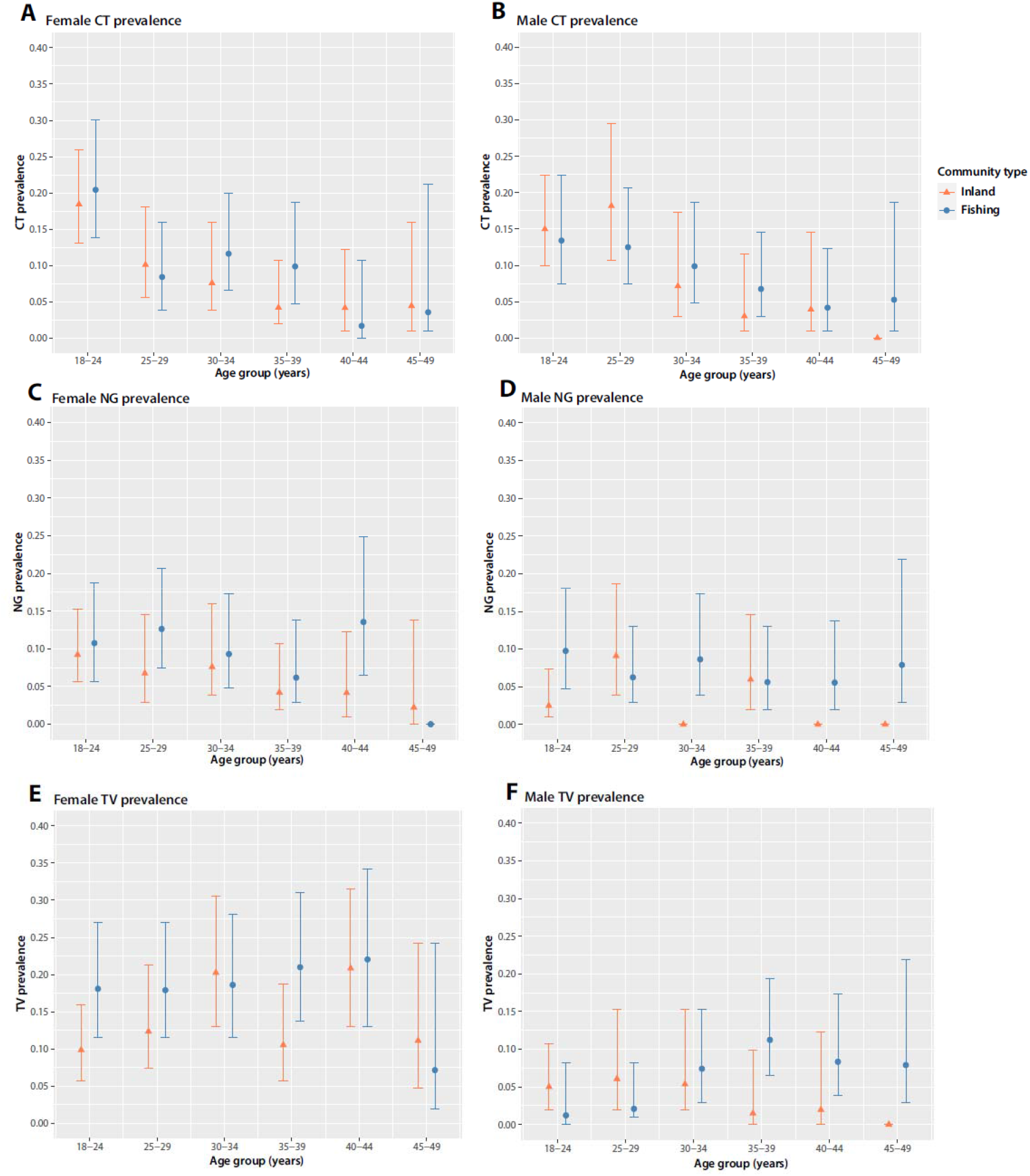
Chlamydia (CT), gonorrhea (NG), and trichomoniasis (TV) prevalence by age, gender, and community (fishing vs. inland).

Gonorrhea was diagnosed in 122 study participants (6.7% prevalence; 95%CI: 5.7-8.0%), including in 5.0% (n=46/919) of inland community participants and 8.4% (n=76/906) of fishing community participants. In contrast to chlamydia, gonorrhea prevalence was significantly higher among HIV-positive compared to HIV-negative participants (10.3 vs. 5.4%; adjPRR=1.84, 95%CI: 1.25-2.72) (Supplemental Table 2), with the exception of women in the fishing community where gonorrhea prevalence was similar irrespective of HIV serostatus (Table 2). There were no clear trends in gonorrhea prevalence by age among men or women (Figure 2C and D).

Overall, prevalence of trichomonas was 11% (95%CI: 9.5-12%). Disease burden was significantly higher among women compared to men (15.9 vs. 5.0%; adjPRR=4.01; 95%CI: 2.36-6.85) and among HIV-positive versus HIV-negative persons (17.0 vs. 8.5%; adjPRR=1.59, 95%CI: 1.18 -2.13) (Supplemental Table 3, Figure 2E and F). In both fishing and inland communities, trichomonas prevalence exceeded 20% among HIV-positive women (Table 2).

HSV-2 seropositivity was highly prevalent in both communities (43% inland, 63% fishing), and was strongly associated with HIV serostatus (Table 2). Women had higher prevalence of HSV-2 than men (63% vs. 43%) irrespective of community or HIV serostatus (adjPRR=1.54; 95%CI: 1.35-1.74). Among the youngest female participants, 18-24 years of age, HSV-2 seroprevalence was 24% (95%CI: 18-32%) in the inland community and 46% (95%CI: 36-56%) in the fishing community. Age-specific seroprevalence increased rapidly among women through their late twenties peaking thereafter in their early thirties. In contrast, male HSV-2 seroprevalence continued to increase steadily through their forties (Figure 3A and B).

**Figure 3.**
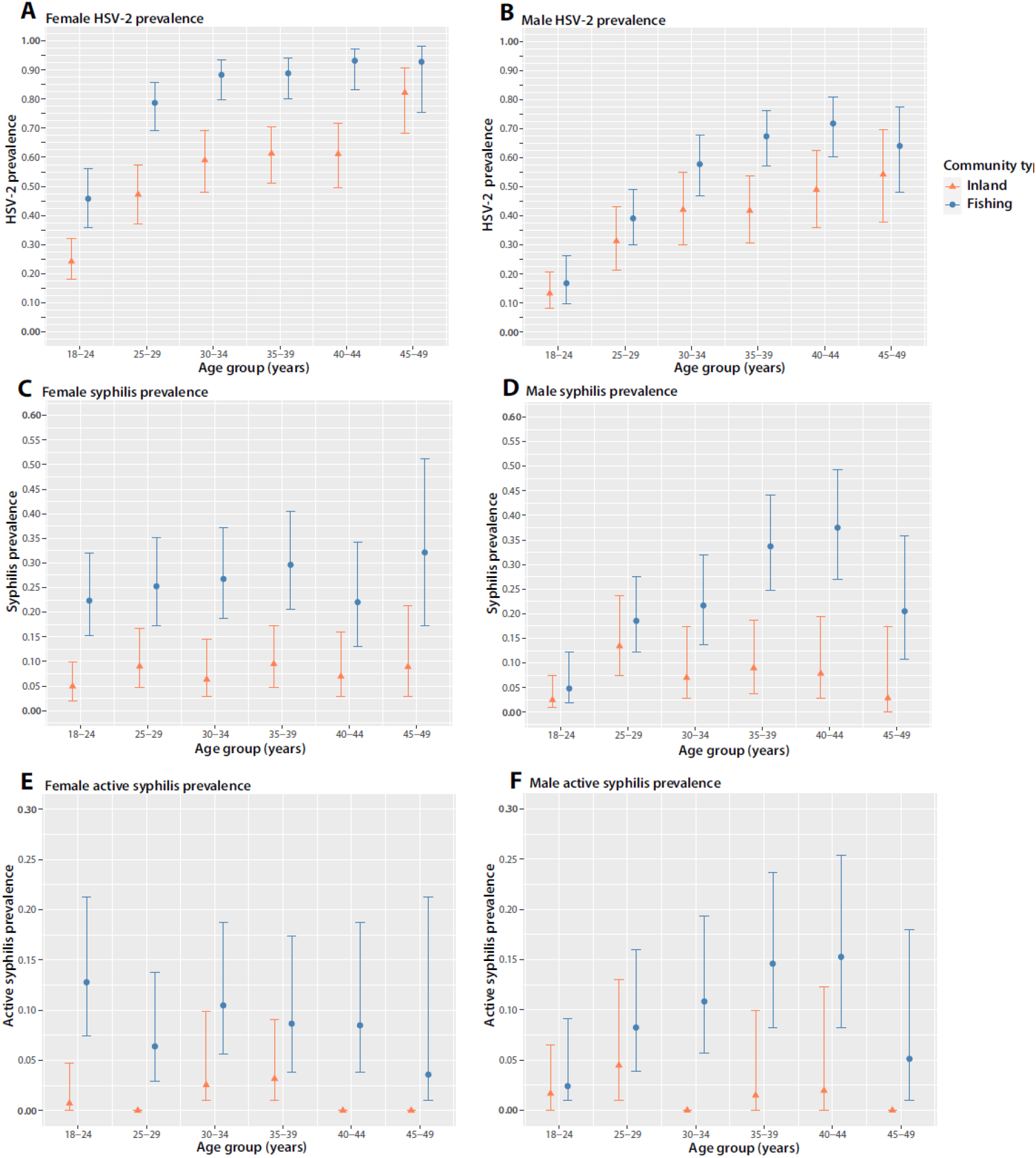
Chlamydia (CT), gonorrhea (NG), and trichomoniasis (TV) prevalence by age, gender, and community (fishing vs. inland).

Syphilis seroprevalence was 7.1% (n=65/919; 95%CI: 5.6-8.9%) among inland residents and 24% (n=219/906; 95%CI: 22-27%) among fishing community residents, with similar burden regardless of gender in both communities. Syphilis seropositivity was strongly and positively associated with HIV serostatus (32.2% vs. 9.4%; adjPRR=2.26; 95%CI: 1.79-2.84), and generally increased with age (Figure 3C and D). Active syphilis infection was far more common among fishing compared to inland community residents (9.4 vs 1.4%; adjPRR=5.17; 95%CI: 2.45-10.91) (Supplemental Table 4). While active syphilis was not associated with female age, its prevalence steadily increased with age among men through their early forties (Figure 3E and F). Notably, 17% (95%CI: 12-24%) of HIV-positive men residing in the fishing community had active syphilis infection.

### Combination HIV treatment and prevention and risk factors for curable STIs

We did not find any statistically significant associations between HIV viral load suppression status or history of PrEP use with either chlamydia, gonorrhea, trichomonas, or active syphilis (Supplemental Tables 1-4). While male circumcision status was associated with significantly reduced prevalence of trichomonas (adjPRR=0.52; 95%CI: 0.29-0.94; Supplemental Table 3), there was substantially higher prevalence of chlamydia among circumcised compared to uncircumcised men (adjPRR=1.78, 95%CI: 1.13-2.79; Supplemental Table 1).

Individuals with higher numbers of sexual partners tended to have higher prevalence of chlamydia, gonorrhea, and active syphilis, but not trichomonas (Supplemental Tables 1-4). For example, chlamydia prevalence was 16% among participant’s reporting ≥ 4 sexual partners in the last year versus 9.2% among those reporting only one partner (adjPRR=1.70; 95%CI: 0.93-3.12). Individuals who were previously married had significantly higher burdens of chlamydia (adjPRR=1.66; 95%CI: 1.17-2.37) and active syphilis (adjPRR=1.84; 95CI: 1.18-2.85) compared to currently married persons.

### STI burden among pregnant women

Of the 107 pregnant women in this study, 31% (n=33) were diagnosed with chlamydia (n=10), gonorrhea (n=7), trichomonas (n=18), or active syphilis (n=3). Overall, burden of curable STIs between women by pregnancy status (adjPRR=0.97, 95%CI: 0.73-1.30). There were five pregnant women who had two curable STIs, including one woman with chlamydia and active syphilis infection: the four other women all had co-infections with trichomonas. All three cases of active syphilis were found among pregnant women residing in the fishing community. There were 24 HIV-positive pregnant women, all of whom, but one, were HIV virally suppressed. Compared to HIV-negative women, HIV-positive women were significantly more likely to have a curable STI (54% vs. 24%, adjPRR=1.87, 95%CI: 1.08-3.23).

### HIV incidence and STIs

There were 876 STIPS participants who tested HIV negative at their previous RCCS study visit, contributing a total of 1,746 person-years of follow-up to the HIV incidence cohort in the two communities. Of these participants, 12 HIV seroconverted for an overall HIV incidence of 0.69 per 100 person-years. HIV incidence was significantly higher in the fishing (1.3 per 100 person-years; n=8 cases/636 person-years) compared to inland community (0.36 per 100 person-years; n=4 cases/1110 person-years; IRR=3.49, 95%CI=1.10-13.1). Gonorrhea and syphilis were more common among HIV incident cases at time of diagnosis, and most (n=10/12) HIV incident cases had HSV-2 antibodies (Table 3).

**Table 3.** HIV incidence and presence of STIs at time of HIV diagnosis

|  | STI positive |  | STI negative |  | IRR (95%CI) | p-value |
| --- | --- | --- | --- | --- | --- | --- |
|  | HIV incident cases/person-years | HIV incidence rate per 100 person-years | HIV incident cases/person-years | HIV incidence rate per 100 person-years |  |  |
| Chlamydia | 0/162.8 | 0 | 12/1578 | 0.76 | - | - |
| Gonorrhea | 2/69.9 | 2.86 | 10/1666.7 | 0.60 | 4.77 (0.73-18.1) | 0.044 |
| Trichomoniasis | 0/139.2 | 0 | 12/1597.4 | 0.75 | - | - |
| Syphilis | 3/153.3 | 1.96 | 9/1593.0 | 0.56 | 3.46 (0.77-11.6) | 0.063 |
| Active Syphilis | 1/47.5 | 2.11 | 11/1698.9 | 0.65 | 3.25 (0.18-16.7) | 0.259 |
| HSV-2 | 10/780.5 | 1.28 | 2/957.7 | 0.21 | 6.2 (1.6-40.2) | 0.019 |

IRR=incidence rate ratio

### Evaluation of syndromic management for STIs

Lastly, we evaluated the sensitivity and specificity of using self-reported STI symptoms for diagnosis of chlamydia, gonorrhea, and trichomonas among men and women (Tables 4-5). Overall, sensitivity of the syndromic approach for all three infections was low irrespective of gender. In contrast, specificity tended to vary depending on STI, symptom, and gender. While combinations of clinically indicative symptoms tended to improve sensitivity, they reduced specificity relative to that of any individual symptom.

**Table 4.** Diagnostic sensitivity and specificity of self-reported symptoms for chlamydia, gonorrhea, and trichomoniasis among women.

|  | **Chlamydia** |  | **Gonorrhea** |  | **Trichomonas** |  |
| --- | --- | --- | --- | --- | --- | --- |
|  | **Sensitivity** | **Specificity** | **Sensitivity** | **Specificity** | **Sensitivity** | **Specificity** |
| **Genital ulcer disease** | 5.2 (1.7,11.6) | 93.2 (91.3,94.8) | 13.2 (6.5,22.9) | 93.9 (92.1,95.4) | 6.5 (3.2,11.7) | 93.3 (91.4,95) |
| **Genital discharge** | 29.9 (21.0,40.0) | 75.8 (72.8,78.6) | 39.5 (28.4,51.4) | 76.4 (73.5,79.2) | 32.7 (25.3,40.7) | 76.6 (73.5,79.5) |
| **Thick/abundant smelly vaginal discharge** | 24.7 (16.5,34.5) | 79.2 (76.4,81.9) | 36.8 (26.1,48.7) | 80.2 (77.4,82.7) | 25.5 (18.8,33.2) | 79.6 (76.6,82.3) |
| **Vaginal itching** | 18.6 (11.4,27.7) | 77.0 (74.1,79.8) | 31.6 (21.4,43.3) | 78.2 (75.4,80.9) | 28.1 (21.1,35.9) | 78.6 (75.6,81.3) |
| **Vaginal odor** | 11.3 (5.8,19.4) | 87.9 (85.5,90.0) | 17.1 (9.4,27.5) | 88.4 (86.1,90.4) | 14.4 (9.2,21) | 88.4 (86,90.5) |
| **Frequent urination** | 13.4 (7.3,21.8) | 92.5 (90.5,94.2) | 7.9 (3.0,16.4) | 91.9 (89.9,93.6) | 9.8 (5.6,15.7) | 92.2 (90.2,94) |
| **Painful urination** | 12.4 (6.6,20.6) | 93.2 (91.3,94.8) | 13.2 (6.5,22.9) | 93.1 (91.3,94.7) | 7.2 (3.6,12.5) | 92.6 (90.6,94.3) |
| **Painful sex** | 8.2 (3.6,15.6) | 93.9 (92.1,95.4) | 9.2 (3.8,18.1) | 93.9 (92.1,95.4) | 5.2 (2.3,10) | 93.5 (91.5,95.1) |
| **Bleeding during sex** | 0.0 (0.0,3.7) | 99.4 (98.7,99.8) | 0.0 (0.0,4.7) | 99.4 (98.7,99.8) | 0.0 (0.0,2.4) | 99.4 (98.6,99.8) |
| **Lower abdominal pain** | 22.7 (14.8,32.3) | 81.1 (78.3,83.6) | 26.3 (16.9,37.7) | 81.3 (78.6,83.8) | 20.3 (14.2,27.5) | 80.8 (77.9,83.4) |
| **Genital warts** | 3.1 (0.6,8.8) | 98.0 (96.9,98.9) | 1.3 (0.0,7.1) | 97.9 (96.7,98.7) | 2.6 (0.7,6.6) | 98.0 (96.8,98.9) |
| **Any symptoms** | 74.2 (64.3,82.6) | 30.9 (27.9,34.1) | 78.9 (68.1,87.5) | 31.2 (28.2,34.4) | 73.9 (66.1,80.6) | 31.2 (28,34.5) |
| **Clinically indicative symptoms\*** | 40.2 (30.4,50.7) | 62.6 (59.3,65.8) | 52.6 (40.8,64.2) | 63.6 (60.3,66.8) | 43.8 (35.8,52) | 63.4 (60,66.7) |

\*Combination of self-reported symptoms indicating treatment (discharge or discharge and abdominal pain) according to the 2016 Uganda Clinical Guidelines

**Table 5.** Diagnostic sensitivity and specificity of self-reported symptoms for chlamydia, gonorrhea, and trichomoniasis among men.

|  | Chlamydia |  | Gonorrhea |  | Trichomonas |  |
| --- | --- | --- | --- | --- | --- | --- |
|  | Sensitivity | Specificity | Sensitivity | Specificity | Sensitivity | Specificity |
| **Genital ulcer disease** | 1.3 (0,6.8) | 97.2 (95.7,98.2) | 4.3 (0.5,14.8) | 97.4 (96,98.4) | 2.3 (0.1,12.3) | 97.3 (95.9,98.3) |
| **Genital discharge** | 10.0 (4.4,18.8) | 96.4 (94.8,97.6) | 34.8 (21.4,50.2) | 97.5 (96.2,98.5) | 7.0 (1.5,19.1) | 95.9 (94.3,97.2) |
| **Frequent urination** | 3.8 (0.8,10.6) | 94.7 (92.9,96.2) | 15.2 (6.3,28.9) | 95.4 (93.7,96.7) | 9.3 (2.6,22.1) | 95.1 (93.3,96.4) |
| **Painful urination** | 7.5 (2.8,15.6) | 93.4 (91.4,95) | 28.3 (16.0,43.5) | 94.5 (92.7,96) | 9.3 (2.6,22.1) | 93.4 (91.5,95.1) |
| **Painful sex** | 1.3 (0.0,6.8) | 97.8 (96.5,98.7) | 10.9 (3.6,23.6) | 98.4 (97.3,99.1) | 2.3 (0.1,12.3) | 97.9 (96.7,98.8) |
| **Bleeding during sex** | 0.0 (0.0,4.5) | 99.9 (99.3,100) | 0.0 (0.0,7.7) | 99.9 (99.3,100) | 0.0 (0.0,8.2) | 99.9 (99.3,100) |
| **Lower abdominal pain** | 0.0 (0.0,4.5) | 96 (94.3,97.3) | 6.5 (1.4,17.9) | 96.5 (95,97.7) | 4.7 (0.6,15.8) | 96.4 (94.9,97.6) |
| **Genital warts** | 2.5 (0.3,8.7) | 96.6 (95.1,97.8) | 8.7 (2.4,20.8) | 97 (95.6,98.1) | 2.3 (0.1,12.3) | 96.7 (95.2,97.8) |
| **Any symptoms** | 37.5 (26.9,49) | 66.2 (62.7,69.5) | 73.9 (58.9,85.7) | 68.1 (64.8,71.3) | 30.2 (17.2,46.1) | 65.6 (62.2,68.9) |
| **Clinically indicative symptoms\*** | 10.0 (4.4,18.8) | 96.4 (94.8,97.6) | 34.8 (21.4,50.2) | 97.5 (96.2,98.5) | 7 (1.5,19.1) | 95.9 (94.3,97.2) |

\*Combination of self-reported symptoms indicating treatment (discharge or painful urination) according to the 2016 Uganda Clinical Guidelines

## Discussion

We found high population-level STI burden in Ugandan communities achieving near universal HIV treatment and high VMMC coverage. Approximately 10% of our study population had chlamydia and gonorrhea, and participants residing in the Lake Victoria fishing community had active syphilis levels exceeding 4 times that of the Ugandan population nationally.^22^ Overall, STI infections were significantly more common among HIV-positive persons, including pregnant women for whom STIs can cause severe adverse pregnancy complications.^1^ Age specific patterns of STI prevalence varied by infection, gender, and community setting. Most STIs detected in this study would not have been diagnosed using the syndromic approach. Despite rapid growth of a massive healthcare infrastructure to diagnose and treat HIV across Africa, our results indicate HIV programs have failed to address non-HIV STI epidemics, especially among HIV-positive populations, and that these infections remain neglected diseases.

Numerous studies over the 40 year history of the HIV pandemic have linked STIs to HIV^23^, including the RCCS in which this study was nested ^24^. We also found strong associations between STIs with both prevalent and incident HIV infection. HIV was most likely to co-occur with syphilis and gonorrhea infections followed by trichomonas. While recent data are lacking, syphilis and trichomonas have been strongly associated with HIV acquisition risk across a range of African settings and populations.^25,26^ In addition, we also found that HSV-2 seroprevalence was nearly universal among HIV-positive participants and associated with a 6-fold higher rate of HIV incidence. Unlike gonorrhea and syphilis, chlamydia is thought to generate a partial protective immune response.^27^ So while chlamydia levels were significantly lower among HIV-positive participants in STIPS, this inverse cross-sectional association likely reflects high levels of pre-existing immunity due past infection, rather than a lower risk of HIV acquisition.

While there are reports of rising STI cases in high-income countries,^4,28^ there is limited data on population-level STI trends in low-income settings, including sub-Saharan Africa. STI data collected from the same geographic region in Uganda as the STIPS study during 1994-1995 reported a syphilis antibody prevalence of 10%, female trichomoniasis prevalence of 24%, gonorrhea prevalence of 1.6%, and chlamydia prevalence of 3.1 %.^24^ In comparison, prevalence of these same STIs in the more comparable inland STIPS community was 7.1%, 13%, 5.0% and 9.6%, respectively. Although the testing platforms and study populations differed between these two studies, a crude comparison of data suggests that prevalence of gonorrhea and chlamydia has remained static or increased, while declining for syphilis and trichomonas. Similar findings have been reported for South Africa, where there have been reductions in syphilis, but not gonorrhea or chlamydia.^29^

Of the 107 pregnant women in this study, nearly one third had a curable STI. These findings are in line with results from other recent African studies showing high prevalence of curable STIs among pregnant women, especially among HIV-positive women.^30^ Of note, we found three cases of active syphilis among pregnant women in STIPS. It is unclear if these cases were the result of screening gaps in antenatal programs or due to missed antenatal visits. In 2016-2017, only 43% of Ugandan antenatal clinic attendees were tested for syphilis in Uganda, while more than ∼95% of pregnant women had attended at least one antenatal visit.^31^ Given the potentially severe sequelae of untreated STIs during pregnancy, these results demonstrate opportunity to improve both maternal and newborn health outcomes.

The strong overlap between HIV and STIs observed here and in prior studies underscores that comprehensive HIV and STI programming remains an unaddressed, but urgent public health priority.^32,33^ A major barrier to integrated HIV/STI Care has been a lack of high quality, affordable STI diagnostics easily implementable in low resource settings.^11^ As shown here, the current approach of syndromic management lacks both sensitivity and specificity leading to substantial underdiagnoses of STIs irrespective of sex and the likely gross misallocation of antibiotic treatment. Emerging STI point of care (POC) diagnostics offer new opportunity for comprehensive HIV and STI programming and more prudent distribution of antibiotic treatments.^34^ The SD Bioline HIV/Syphilis Duo test, a dual syphilis and HIV rapid screening test, has been validated in multiple African settings,^35,36^ but is not yet widely integrated into HIV testing programs. Over the last decade, there has been major advancement in the development of POC STI diagnostics for other curable STIs including the OSOM lateral flow test for trichomonas,^37^ the Binx health io test for gonorrhea and chlamydia,^38^ and the Visby test, for gonorrhea, chlamydia and trichomonas ^39^. While these tests theoretically could lead to more timely and improved STI care in sub-Saharan Africa, their effectiveness and implementation still needs rigorous evaluation in low resource settings and cost remains a major barrier to rollout.

Few STIPS participants reported PrEP use, limiting our ability to draw any conclusions on PrEP and STIs in this study. Oral HIV PrEP has been associated with higher risk of STI incidence among MSM.^40^ Increases in risk following PrEP initiation is hypothesized to be due to increases in risky sexual behavior with perceived protection from HIV (i.e., risk compensation), although there is considerable debate about the causal link between oral PrEP use and STIs.^41^ As of 2019, thirty percent of all global PrEP users were in sub-Saharan Africa, many of whom are women, but little is known about the association between STI incidence and PrEP initiation within context of programmatic PrEP rollout regionally.^42^

This study has limitations. First, the study was conducted in two communities in southern Uganda with moderate and extremely HIV burden, and so these results may not be generalizable to other settings, particularly those with lower levels of HIV burden and STI risk factors. However, population-level data on STI prevalence is rare and the data generated here are in line with an earlier population-based study in South Africa, which similarly reported high STI burden.^43^ The OSOM rapid test for trichomonas is not validated for male penile meatal swabs as was collected and tested here, and therefore these results may be underestimates. While we report on STIs among a small number of HIV incident cases, STIs were detected at time of first positive HIV test and may have been acquired during or after HIV acquisition. While we cannot conclude that STIs are risk factors for HIV acquisition from this cross-sectional study, many studies have shown that STIs are associated with HIV acquisition.^23^ Even if STIs are not causally linked to disease acquisition, they serve as important markers of high-risk sexual activity independent of self-reported HIV risk behaviors.

In conclusion, we found extremely high STI burden in this population-based study conducted in two African communities with a high HIV burden and near universal HIV treatment coverage. STIs were particularly elevated among HIV-positive persons, most of whom were engaged in HIV services, including pregnant women. Taken together, these data suggest a missed opportunity to reduce STIs in sub-Saharan Africa. Investment in combination HIV and STI programming has the potential to reduce HIV incidence, while at the same time improving female reproductive and neonatal health. Rollout of POC STI diagnostics will be critical for achieving integrated program success but must be implemented using sustainable, cost-effective approaches.

## Data Availability

All data produced in the present study are available upon reasonable request to the authors

## Acknowledgements

This work was supported by grants from the Johns Hopkins Center for AIDS Research (grant number P30AI094189), grants from the National Institute of Allergy and Infectious Diseases (grant numbers R01AI143333, R01MH115799, K01AI125086); the National Institute of Mental Health (grant number R01MH107275); the Eunice Kennedy Shriver National Institute of Child Health and Human Development (grant number RO1HD091003); the Division of Intramural Research of the National Institute for Allergy and Infectious Diseases; and the President’s Emergency Plan for AIDS Relief through the Centers for Disease Control and Prevention (grant number NU2GGH000817). We thank the personnel at the Office of Cyberinfrastructure and Computational Biology at the National Institute of Allergy and Infectious Diseases for data management support. Additionally, we thank the cohort participants and many staff and investigators who have made this study possible over the years.

## Supplemental Tables

**Supplemental Table 1.** Risk factors for chlamydia.

|  | n/Total | PRR | p-value | adjPRR | p-value |
| --- | --- | --- | --- | --- | --- |
| **Overall** |  |  |  |  |  |
| **Sex** |  |  |  |  |  |
| Female | 97/963 (10.1) | 1.07 (0.81-1.42) | 0.623 | 1.06 (0.69-1.62) | 0.805 |
| Male | 80/852 (9.4) | Ref. | - | Ref. | - |
| **Community type** |  |  |  |  |  |
| Inland | 88/915 (9.6) | Ref. | - | Ref. | ;- |
| Fishing | 89/900 (9.9) | 1.03 (0.78-1.36) | 0.846 | 0.87 (0.63-1.21) | 0.421 |
| **Age** |  |  |  |  |  |
| 15-24 | 74/436 (17.0) | Ref. | - | Ref. | - |
| 25-29 | 41/436 (11.8) | **0.70 (0.49-1.00)** | **0.047** | **0.56 (0.38-0.83)** | **0.004** |
| 30-34 | 28/302 (9.3) | **0.55 (0.36-0.82)** | **0.004** | **0.46 (0.30-0.71)** | **<0.001** |
| 35-39 | 20/332 (6.0) | **0.35 (0.22-0.57)** | **<0.001** | **0.32 (0.19-0.53)** | **<0.001** |
| 40-44 | 9/254 (3.5) | **0.21 (0.11-0.41)** | **<0.001** | **0.20 (0.10-0.41)** | **<0.001** |
| 45-49 | 5/145 (3.4) | **0.20 (0.08-0.49)** | **<0.001** | **0.21 (0.08-0.53)** | **0.001** |
| **Marital Status** |  |  |  |  |  |
| Currently married | 108/1194 (9.0) | Ref. | - | Ref. | - |
| Never married | 23/260 (8.8) | 0.98 (0.65-1.50) | 0.919 | 0.68 (0.40-1.15) | 0.150 |
| Previously married | 46/361 (12.7) | **1.41 (1.02-1.95)** | **0.038** | **1.66 (1.17-2.37)** | **0.005** |
| **Education** |  |  |  |  |  |
| None | 9/133 (6.8) | 0.73 (0.38-1.40) | 9.338 | 0.87 (0.45-1.67) | 0.676 |
| Primary | 106/1136 (9.3) | Ref. | - | Ref. | - |
| Secondary | 36/403 (8.9) | 0.96 (0.67-1.37) | 0.813 | 0.92 (0.64-1.34) | 0.676 |
| Tertiary/Trade School | 26/143 (18.2) | **1.95 (1.32-2.88)** | **<0.001** | 1.46 (0.97-2.21) | 0.071 |
| **Occupation** |  |  |  |  |  |
| Agriculture | 34/515 (6.6) | Ref. | - | Ref. | - |
| Transport (boda boda/mechanic) | 10/54 (18.5) | 2.81 (1.47-5.36) | 0.0018 | 1.87 (0.96-3.66) | 0.068 |
| Fishing | 22/287 (7.7) | 1.16 (0.69-1.95) | 0.571 | 1.15 (0.6-2.18) | 0.678 |
| Government/clerical/teaching | 9/71 (12.7) | 1.92 (0.96-3.83) | 0.064 | 1.63 (0.82-3.22) | 0.161 |
| Hair dresser | 9/53 (17.0) | **2.57 (1.31-5.07)** | **0.006** | 1.49 (0.72-3.07) | 0.282 |
| Housework | 16/111 (14.4) | **2.18 (1.25-3.81)** | **0.006** | 1.71 (0.94-3.11) | 0.077 |
| Student | 1/52 (1.9) | 0.29 (0.04-2.08) | 0.219 | 0.47 (0.06-3.96) | 0.488 |
| Trader/shop keeper | 31/331 (9.4) | 1.42 (0.89-2.26) | 0.142 | 1.35 (0.83-2.20) | 0.220 |
| Bartender/waitress | 16/92 (17.4) | **2.63 (1.52-4.57)** | **<0.001** | **2.29 (1.28-4.11)** | **0.005** |
| Other | 29/249 (11.6) | **1.76 (1.10-2.83)** | **0.018** | 1.55 (0.93-2.59) | 0.091 |
| **Sex partners in the last year** |  |  |  |  |  |
| None | 4/178 (2.2) | **0.25 (0.09-0.66)** | **0.005** | **0.31 (0.1-0.93)** | **0.036** |
| One | 96/1049 (9.2) | Ref. | - | Ref. | - |
| Two | 39/338 (11.5) | **1.26 (0.89-1.79)** | **0.196** | 1.27 (0.85-1.88) | 0.240 |
| Three | 21/144 (14.6) | **1.59 (1.03-2.47)** | **0.037** | 1.49 (0.9-2.47) | 0.122 |
| Four or more | 17/106 (16.0) | **1.75 (1.09-2.82)** | **0.021** | 1.70 (0.93-3.12) | 0.085 |
| **HIV prevalence** |  |  |  |  |  |
| HIV negative | 146/1329 (10.9) | Ref. | - |  |  |
| HIV positive | 32/486 (6.6) | **0.60 (0.42-0.87)** | **0.007** | 0.75 (0.52-1.09) | 0.1367 |
| **Viral load suppression status\*** |  |  |  |  |  |
| ≤1000 copies/ml | 31/440 (7.0) | Ref. | - | Ref. | - |
| >1000 copies/ml | 1/46 (2.2) | 0.31 (0.04-2.1) | 0.242 | 0.26 | 0.154 |
| **Male circumcision status\*\*** |  |  |  |  |  |
| Uncircumcised | 27/424 (6.3) | Ref. | - | Ref. | - |
| Circumcised | 53/428 (11.9) | **1.94 (1.25-3.03)** | **0.003** | **1.78 (1.13-2.79)** | **0.012** |
| **Ever used PrEP†** |  |  |  |  |  |
| No | 142/1284 (11.1) | Ref. | - | Ref. | - |
| Yes | 3/43 (7.0) | 0.63 (0.21-1.90) | 0.412 | 0.70 (0.23-2.19) | 0.545 |
| **Currently pregnant‡** |  |  |  |  |  |
| No | 87/857 (10.2) | Ref. | - | Ref. | - |
| Yes | 10/106 (9.4) | 0.93 (0.50-1.73) | 0.817 | 0.76 (0.41-1.41) | 0.386 |

PRR=prevalence risk ratio; adjPRR=adjusted prevalence risk ratio; Primary adjusted model included all covariates, with exception viral load suppression status, male circumcision status, PrEP use, and pregnancy status. Adjustment for these latter four variables was done in separate stratified analyses by sex and HIV status. \*Viral load suppression status only reported for HIV-positive participants; adjusted analyses were restricted to HIV-positive participants only and included all covariates except for HIV status, PrEP use, and pregnancy status. \*\* Analyses were restricted to men only. Adjusted analysis included all covariates except for viral load suppression status, PrEP use, and pregnancy status †There was missing data on PrEP use for 2 participants; analyses restricted to HIV negative participants only. Adjusted analysis included all covariates except for HIV status, viral load suppression status, and pregnancy status; ‡ Analyses were restricted to women only. Adjusted analysis included all covariates except for viral load suppression status, PrEP use, and male circumcision status.

**Supplemental Table 2.** Risk factors for gonorrhea.

|  | N/Total (%) | PRR | p-value | adjPRR | p-value |
| --- | --- | --- | --- | --- | --- |
| **Sex** |  |  |  |  |  |
| Female | 76/963 (7.9) | **1.46 (1.03-1.46)** | **0.036** | **2.09 (1.14-3.82)** | **0.017** |
| Male | 46/852 (5.4) | Ref. | - | Ref. | - |
| **Community type** |  |  |  |  |  |
| Inland | 46/915 (5.0) | Ref. | - | Ref. | - |
| Fishing | 76/900 (8.4) | **1.68 (1.18-2.39)** | **0.004** | 1.05 (0.67-1.66) | 0.821 |
| **Age** |  |  |  |  |  |
| 15-24 | 34/436 (7.8) | Ref. | - |  |  |
| 25-29 | 30/346 (8.7) | 1.11 (0.69-1.78) | 0.659 | 0.85 (0.52-1.42) | 0.544 |
| 30-34 | 21/302 (7.0) | 0.89 (0.53-1.51) | 0.668 | 0.60 (0.34-1.04) | 0.067 |
| 35-39 | 18/332 (5.4) | 0.70 (0.40-1.21) | 0.198 | **0.54 (0.30-1.00)** | **0.051** |
| 40-44 | 16/254 (5.9) | 0.76 (0.42-1.36) | 0.354 | 0.60 (0.31-1.17) | 0.135 |
| 45-49 | 4/145 (2.8) | 0.35 (0.13-0.98) | 0.046 | **0.36 (0.13-1.00)** | **0.050** |
| **Marital Status** |  |  |  |  |  |
| Currently married | 88/1194 (7.4) | Ref. | - |  |  |
| Never married | 14/260 (5.4) | 0.73 (0.42-1.26) | 0.261 | 1.09 (0.61-1.97) | 0.770 |
| Previously married | 20/361 (5.5) | 0.75 (0.47-1.20) | 0.234 | 0.70 (0.43-1.16) | 0.164 |
| **Education** |  |  |  |  |  |
| None | 8/133 (6.0) | 0.76 (0.38-1.53) | 0.441 | 0.72 (0.36-1.48) | 0.376 |
| Primary | 90/1136 (7.9) | Ref. | - |  |  |
| Secondary | 13/403 (3.2) | **0.41 (0.23-0.72)** | **0.002** | **0.48 (0.28-0.84)** | **0.010** |
| Tertiary/Trade School | 11/143 (7.7) | 0.97 (0.53-1.77) | 0.923 | 1.15 (0.58-2.3) | 0.692 |
| **Occupation** |  |  |  |  |  |
| Agriculture | 27/515 (5.2) | Ref. | - | Ref. | - |
| Transport (boda boda/mechanic) | 2/54 (3.7) | 0.71 (0.17-2.89) | 0.629 | 0.67 (0.16-2.72) | 0.570 |
| Fishing | 23/287 (8.0) | 1.53 (0.89-2.62) | 0.121 | 1.51 (0.73-3.1) | 0.264 |
| Government/clerical/teaching | 1/71 (1.4) | 0.27 (0.04-1.95) | 0.193 | 0.27 (0.03-2.22) | 0.224 |
| Hair dresser | 4/53 (7.5) | 1.44 (0.52-3.96) | 0.480 | 1.01 (0.32-3.15) | 0.988 |
| Housework | 11/111 (9.9) | 1.89 (0.97-3.70) | 0.063 | 1.05 (0.51-2.18) | 0.895 |
| Student | 0/52 (0) | - | -- | **0 (0-0)** | **<0.001** |
| Trader/shop keeper | 29/331 (8.8) | 1.67 (1.01-2.77) | 0.047 | 1.41 (0.82-2.43) | 0.214 |
| Bartender/waitress | 11/92 (12.0) | 2.28 (1.17-4.43) | 0.015 | 1.61 (0.76-3.43) | 0.218 |
| Other | 14/249 (5.6) | 1.07 (0.57-2.01) | 0.827 | 0.89 (0.46-1.73) | 0.735 |
| **Sex partners in the last year** |  |  |  |  |  |
| None | 2/178 (1.1) | 0.17 (0.04-0.69) | 0.013 | 0.27 (0.06-1.12) | 0.072 |
| One | 69/1049 (6.6) | Ref. | - |  |  |
| Two | 25/338 (7.4.) | 1.12 (0.72-1.75) | 0.602 | 1.45 (0.89-2.38) | 0.138 |
| Three | **16/144 (11.1)** | **1.69 (1.01-2.83)** | **0.046** | **2.18 (1.22-3.9)** | **0.008** |
| Four or more | 10/106 (9.4) | 1.43 (0.76-2.70) | 0.264 | **2.06 (1.01-4.21)** | **0.047** |
| **HIV prevalence** |  |  |  |  |  |
| HIV negative | 72/139 (5.4) | Ref. | - | Ref. | - |
| HIV positive | 50/486 (10.3) | **1.90 (1.34-2.68)** | **<0.001** | **1.84 (1.25-2.72)** | **0.002** |
| **Viral load suppression status\*** |  |  |  |  |  |
| ≤1000 copies/ml | 44/440 (10.0) | Ref. | - | Ref. | - |
| >1000 copies/ml | 6/46 (13.0) | 1.30 (0.59-2.89) | 0.514 | 1.20 (0.53-2.74) | 0.660 |
| **Male circumcision status\*\*** |  |  |  |  |  |
| Uncircumcised | 24/424 (6.4) | Ref. | - | Ref. | - |
| Circumcised | 19/428 (4.4) | 0.70 (0.39-1.23) | 0.216 | 0.84 (0.45-1.59) | 0.600 |
| **Ever used PrEP†** |  |  |  |  |  |
| No | 70/1284 (5.5) | Ref. |  | Ref. | - |
| Yes | 2/43 (4.7) | 0.85 (0.22-3.37) | 0.821 | 0.74 (0.21-2.63) | 0.640 |
| **Currently pregnant‡** |  |  |  |  |  |
| No | 69/857 (8.1) | Ref. | - | Ref. | - |
| Yes | 7/106 (6.6) | 0.82 (0.39-1.74) | 0.605 | 0.74 (0.36-1.54) | 0.425 |

PRR=prevalence risk ratio; adjPRR=adjusted prevalence risk ratio; Primary adjusted model included all covariates, with exception viral load suppression status, male circumcision status, PrEP use, and pregnancy status. Adjustment for these latter four variables was done in separate stratified analyses by sex and HIV status. \*Viral load suppression status only reported for HIV-positive participants; adjusted analyses were restricted to HIV-positive participants only and included all covariates except for HIV status, PrEP use, and pregnancy status. \*\* Analyses were restricted to men only. Adjusted analysis included all covariates except for viral load suppression status, PrEP use, and pregnancy status †There was missing data on PrEP use for 2 participants; analyses restricted to HIV negative participants only. Adjusted analysis included all covariates except for HIV status, viral load suppression status, and pregnancy status; ‡ Analyses were restricted to women only. Adjusted analysis included all covariates except for viral load suppression status, PrEP use, and male circumcision status.

**Supplemental Table 3.** Risk factors for trichomonas.

|  | N/Total (%) | PRR | p-value | adjPRR | p-value |
| --- | --- | --- | --- | --- | --- |
| **Gender** |  |  |  |  |  |
| Female | 153/965 (15.9) | **3.14 (2.27-4.35)** | **<0.001** | **4.01 (2.35-6.85)** | **<0.001** |
| Male | 43/852 (5.0) | Ref. | - | Ref. | - |
| **Community type** |  |  |  |  |  |
| Inland | 86/916 (9.4) | Ref. | - | Ref. | - |
| Fishing | 110/901 (12.2) | 1.30 (1.00-1.70) | 0.054 | 0.92 (0.64-1.32) | 0.647 |
| **Age** |  |  |  |  |  |
| 15-24 | 38/438 (8.7) | Ref. | - | Ref. | - |
| 25-29 | 34/346 (9.8) | 1.13 (0.73-1.76) | 0.579 | 0.84 (0.53-1.31) | 0.435 |
| 30-34 | 41/302 (13.6) | **1.56 (1.03-2.37)** | **0.035** | 1.02 (0.66-1.59) | 0.915 |
| 35-39 | 38/332 (11.4) | 1.32 (0.86-2.02) | 0.203 | 0.91 (0.59-1.42) | 0.687 |
| 40-44 | 35/254 (13.8) | 1.59 (1.03-2.45) | 0.036 | 1.12 (0.71-1.76) | 0.638 |
| 45-49 | 10/145 (6.9) | 0.79 (0.41-1.55) | 0.502 | 0.62 (0.31-1.22) | 0.166 |
| **Marital Status** |  |  |  |  |  |
| Currently married | 128/1196 (10.7) | Ref. | - | Ref. | - |
| Never married | 13/260 (5.0) | **0.47 (0.27-0.81)** | **0.007** | 0.85 (0.48-1.52) | 0.582 |
| Previously married | 55/361 (15.2) | **1.42 (1.06-1.91)** | **0.018** | 1.29 (0.94-1.78) | 0.114 |
| **Education** |  |  |  |  |  |
| None | 22/133 (16.5) | 1.43 (0.94-2.16) | 0.093 | 1.16 (0.76-1.77) | 0.498 |
| Primary | 132/1138 (11.6) | Ref. | - | Ref. | - |
| Secondary | 29/403 (7.2) | **0.62 (0.42-0.91)** | **0.015** | 0.68 (0.46-1.02) | 0.062 |
| Tertiary/Trade School | 13/143 (9.1) | 0.78 (0.46-1.35) | 0.379 | 0.76 (0.4-1.42) | 0.386 |
| **Occupation** |  |  |  |  |  |
| Agriculture | 53/516 (10.3) | Ref. | - | Ref. | - |
| Transport (boda boda/mechanic) | 3/54 (5.4) | 0.54 (0.17-1.67) | 0.286 | 1.56 (0.46-5.22) | 0.474 |
| Fishing | 22/287 (7.7) | 0.75 (0.46-1.20) | 0.228 | 1.65 (0.81-3.38) | 0.167 |
| Government/clerical/teaching | 7/71 (9.9) | 0.96 (0.45-2.03) | 0.915 | 1.29 (0.56-2.97) | 0.545 |
| Hair dresser | 4/53 (7.5) | 0.73 (0.28-1.95) | 0.536 | 0.71 (0.25-2.07) | 0.535 |
| Housework | 20/111 (18.0) | **1.75 (1.09-2.81)** | **0.020** | 1.11 (0.64-1.93) | 0.701 |
| Student | 1/52 (1.9) | 0.19 (0.03-1.33) | 0.093 | 0.56 (0.07-4.74) | 0.593 |
| Trader/shop keeper | 44/332 (13.3) | 1.29 (0.89-1.88) | 0.183 | 1.22 (0.8-1.84) | 0.357 |
| Bartender/waitress | 21/92 (22.8) | **2.22 (1.41-3.50)** | **0.001** | 1.4 (0.84-2.34) | 0.200 |
| Other | 21/249 (8.4) | 0.82 (0.51-1.33) | 0.423 | 0.86 (0.51-1.44) | 0.559 |
| **Sex partners in the last year** |  |  |  |  |  |
| None | 10/178 (5.6) | **0.45 (0.24-0.83)** | **0.011** |  |  |
| One | 142/1051 (12.6) | Ref. | - | Ref. | - |
| Two | 33/338 (9.8) | 0.78 (0.54-1.12) | 0.172 | 1.2 (0.82-1.77) | 0.346 |
| Three | 9/144 (6.2) | 0.50 (0.26-0.96) | 0.036 | 0.95 (0.48-1.87) | 0.871 |
| Four or more | 12/106 (11.3) | 0.90 (0.52-1.57) | 0.714 | 1.69 (0.9-3.19) | 0.105 |
| **HIV serostatus** |  |  |  |  |  |
| HIV negative | 113/1329 (8.5) | Ref. | - | Ref. | - |
| HIV positive | 83/488 (17.0) | **2.0 (1.54-2.6)** | **<0.001** | **1.59 (1.18-2.13)** | **0.002** |
| **Viral load suppression status\*** |  |  |  |  |  |
| ≤1000 copies/ml | 77/442 (17.4) | Ref. | - | Ref. | - |
| >1000 copies/ml | 6/46 (13.0) | 0.75 (0.35-1.62) | 0.463 | 0.85 (0.40-1.83) | 0.675 |
| **Male circumcision status\*\*** |  |  |  |  |  |
| Uncircumcised | 29/424 (6.8) | Ref. | - | Ref. | - |
| Circumcised | 14/428 (3.3) | **0.48 (0.26-0.89)** | **0.020** | **0.52 (0.29-0.94)** | **0.035** |
| **Ever used PrEP†** |  |  |  |  |  |
| No | 108/1274 (8.4) | Ref. | - | Ref. | - |
| Yes | 5/43 (11.6) | 1.38 (0.59-3.21) | 0.452 | 1.21 (0.52-2.82) | 0.667 |
| **Currently pregnant‡** |  |  |  |  |  |
| No | 135/858 (15.7) | Ref. | - | Ref. | - |
| Yes | 18/107 (16.8) | 1.07 (0.68-1.67) | 0.770 | 1.06 (0.68-1.67) | 0.785 |

PRR=prevalence risk ratio; adjPRR=adjusted prevalence risk ratio; Primary adjusted model included all covariates, with exception viral load suppression status, male circumcision status, PrEP use, and pregnancy status. Adjustment for these latter four variables was done in separate stratified analyses by sex and HIV status. \*Viral load suppression status only reported for HIV-positive participants; adjusted analyses were restricted to HIV-positive participants only and included all covariates except for HIV status, PrEP use, and pregnancy status. \*\* Analyses were restricted to men only. Adjusted analysis included all covariates except for viral load suppression status, PrEP use, and pregnancy status †There was missing data on PrEP use for 2 participants; analyses restricted to HIV negative participants only. Adjusted analysis included all covariates except for HIV status, viral load suppression status, and pregnancy status; ‡ Analyses were restricted to women only. Adjusted analysis included all covariates except for viral load suppression status, PrEP use, and male circumcision status.

**Supplemental Table 4.** Risk factors for active syphilis.

|  | N/Total (%) | PRR | p-value | adjPRR | p-value |
| --- | --- | --- | --- | --- | --- |
| **Sex** |  |  |  |  |  |
| Female | 46/964 (4.8) | 0.79 (0.54-1.16) | 0.229 | 1.18 (0.55-2.52) | 0.669 |
| Male | 52/860 (6.0) | Ref. | - | Ref. | - |
| **Community type** |  |  |  |  |  |
| Inland | 13/919 (1.4) | Ref. | - | Ref. | - |
| Fishing | 85/905 (9.4) | **6.64 (3.73-11.82)** | **<0.001** | **5.17 (2.45-10.91)** | **<0.001** |
| **Age** |  |  |  |  |  |
| 15-24 | 17/439 (3.9) | Ref. | - | Ref. | - |
| 25-29 | 17/347 (4.9) | 1.27 (0.66-2.44) | 0.580 | 0.84 (0.43-1.64) | 0.613 |
| 30-34 | 20/305 (6.6) | **1.69 (0.90-3.18)** | **0.035** | 0.99 (0.5-1.98) | 0.984 |
| 35-39 | 24/332 (7.2) | 1.87 (1.02-3.42) | 0.203 | 1.22 (0.63-2.39) | 0.556 |
| 40-44 | 17/254 (6.7) | **1.73 (0.90-3.32)** | **0.036** | 0.98 (0.46-2.07) | 0.954 |
| 45-49 | 3/147 (2.0) | 0.53 (0.16-1.77) | 0.502 | 0.37 (0.11-1.28) | 0.117 |
| **Marital Status** |  |  |  |  |  |
| Currently married | 57/1199 (4.8) | Ref. | - | Ref. | - |
| Never married | 8/262 (3.1) | 0.64 (0.31-1.33) | 0.233 | 1.33 (0.58-3.09) | 0.502 |
| Previously married | 33/363 (9.1) | **1.91 (1.27-2.89)** | **0.002** | **1.84 (1.18-2.85)** | **0.007** |
| **Education** |  |  |  |  |  |
| None | 11/133 (8.3) | .125 (0.69-2.31) | 0.093 | 0.91 (0.52-1.6) | 0.745 |
| Primary | 75/1143 (6.6) | Ref. | - | Ref. | - |
| Secondary | .7/405 (1.7) | **0.26 (0.12-0.57)** | **0.001** | **0.48 (0.23-1.03)** | **0.061** |
| Tertiary/Trade School | 5/143 (3.5) | 0.53 (0.22-1.30) | 0.165 | 1 (0.42-2.39) | 0.999 |
| **Occupation** |  |  |  |  |  |
| Agriculture | 14/516 (2.7) | Ref. | - | Ref. | - |
| Transport (boda boda/mechanic) | 3/54 (5.6) | 2.05 (0.61-6.90) | 0.248 | 0.97 (0.28-3.31) | 0.963 |
| Fishing | 36/289 (12.5) | **4.59 (2.52-8.37)** | **<0.001** | 0.92 (0.39-2.15) | 0.842 |
| Government/clerical/teaching | 0/71 (0) | - | - | - | - |
| Hair dresser | 1/53 (1.9) | 0.70 (0.09-5.18) | 0.723 | 0.34 (0.05-2.42) | 0.278 |
| Housework | 17/110 (15.5) | **5.70 (2.90-11.21)** | **<0.001** | 1.26 (0.55-2.89) | 0.592 |
| Student | 0/52 (0) | - | - | - | - |
| Trader/shop keeper | 11/335 (3.3) | 1.21 (0.56-2.63) | 0.631 | **0.39 (0.16-0.95)** | **0.039** |
| Bartender/waitress | 4/92 (4.3) | 1.60 (0.54-4.76) | 0 | 0.35 (0.11-1.13) | 0.079 |
| Other | 12/252 | 1.76 (0.82-3.74) | 0.145 | 0.49 (0.2-1.17) | 0.108 |
| **Sex partners in the last year** |  |  |  |  |  |
| None | 4/179 (2.2) | 0.49 (0.18-1.35) | 0.167 | 0.64 (0.24-1.71) | 0.372 |
| One | 48/1055 (4.5) | Ref. | - | Ref. | - |
| Two | 22/339 (6.5) | 1.43 (0.87-2.33) | 0.156 | 1.22 (0.7-2.1) | 0.482 |
| Three | 13/145 (9.0) | 1.97 (1.09-3.55) | 0.024 | 1.48 (0.77-2.83) | 0.238 |
| Four or more | 11/106 (10.4) | 2.28 (1.22-4.26) | 0.010 | 1.16 (0.59-2.28) | 0.660 |
| **HIV prevalence** |  |  |  |  |  |
| HIV negative | 47/1334 (3.5) | Ref. | - | Ref. | - |
| HIV positive | 51/490 (10.4) | **2.95 (2.02-4.33)** | **<0.0001** | **1.77 (1.15-2.72)** | **0.009** |
| **Viral load suppression status\*** |  |  |  |  |  |
| ≤1000 copies/ml | 48/444 (10.8) | Ref. | - | Ref. | - |
| >1000 copies/ml | 3/46 (6.5) | 0.60 (0.20-1.86) | 0.379 | 0.52 (0.15-1.81) | 0.301 |
| **Male circumcision status\*\*** |  |  |  |  |  |
| Uncircumcised | 24/388 (6.2) | Ref. | - | Ref. | - |
| Circumcised | 28/472 (5.9) | 0.96 (0.57-1.63) | 0.877 | 1.22 (0.71-2.09) | 0.462 |
| **Ever used PrEP†** |  |  |  |  |  |
| No | 44/1289 (3.4) | Ref. | - | Ref. | - |
| Yes | 3/43 (7.0) | 2.04 (0.66-6.32) | 0.215 | 1.03 (0.34-3.11) | 0.955 |
| **Currently pregnant‡** |  |  |  |  |  |
| No | 43/858 | Ref. | - | Ref. | - |
| Yes | 3/106 | 0.56 (0.18-1.79) | 0.331 | 0.61 (0.20-1.88) | 0.390 |

PRR=prevalence risk ratio; adjPRR=adjusted prevalence risk ratio; Primary adjusted model included all covariates, with exception viral load suppression status, male circumcision status, PrEP use, and pregnancy status. Adjustment for these latter four variables was done in separate stratified analyses by sex and HIV status. \*Viral load suppression status only reported for HIV-positive participants; adjusted analyses were restricted to HIV-positive participants only and included all covariates except for HIV status, PrEP use, and pregnancy status. \*\* Analyses were restricted to men only. Adjusted analysis included all covariates except for viral load suppression status, PrEP use, and pregnancy status. †There was missing data on PrEP use for 2 participants; analyses restricted to HIV negative participants only. Adjusted analysis included all covariates except for HIV status, viral load suppression status, and pregnancy status; ‡ Analyses were restricted to women only. Adjusted analysis included all covariates except for viral load suppression status, PrEP use, and male circumcision status.

